# How COVID-19 pandemic impacted the students and staff physical activity? A study in a Southern Brazilian University

**DOI:** 10.1101/2022.11.23.22280287

**Authors:** Rafaela Costa Martins, Luiza Isnardi Cardoso Ricardo, Inácio Crochemore-Silva, Flávio Fernando Demarco, Tiago N Munhoz, Mateus L Levandowski, Mariana Gonzalez Cademartori, Helena Silveira Schuch, Pedro Curi Hallal

## Abstract

Physical activity and its positive effects on coronavirus have been extensively discussed in the literature. However, there is still lack of evidence on the effects of the coronavirus pandemic on the health-related behaviors of the Brazilian university community. The aim of the present study is to describe physical activity practice during the coronavirus pandemic among students and staff of a southern Brazilian university, as well as its association with sociodemographic characteristics. This was a self-administered web-based cross-sectional study, carried out among a southern Brazilian community. The main outcome for this study was leisure time physical activity during the coronavirus pandemic. Considering only leisure-time, 21.0% and 24.0% of the students and staff, respectively, reported achieving physical activity recommendations (≥150 minutes per week). There was a decline of more than 15 percentage points in physical activity practice comparing pre- and during the pandemic, and those following the protocols of staying at home presented lower levels of leisure-time physical activity. Physical activity practice was mainly performed at home and without any professional help. Leisure-time physical activity prevalence during the pandemic was relatively low among students and staff, and participants that followed staying at home protocols presented lower levels of physical activity.

## Introduction

The COVID-19 pandemic rapidly spread in 2020 and remains impacting the social and health dynamics worldwide. In response, the World Health Organization (WHO) published recommendations of protective measures to limit the circulation of the virus in the population (World Health Organization, 2020), with strong emphasis on social distancing and stay at home orientation. To minimally comply with these measures, a series of lifestyle changes were necessary, potentially impacting negatively on some health-related behaviors (Zvolensky et al. 2020).

Physical activity (PA) and its positive effects on COVID-19 have been extensively discussed in the literature. PA is related to improvements in the immune system (Nieman et al. 2019) and is associated with risk factors for COVID-19 severity, such as obesity, cardiovascular diseases, and diabetes (Lee et al. 2012). In addition, the progression of the disease is much worse in inactive individuals, since they have greater risk of hospitalizations, admission in intensive care units, and deaths due to COVID-19 (Sallis et al. 2016). Along with the biological effects, positive sociocultural effects can also be expected from an active lifestyle, helping decrease the mental health burden related to the COVID-19 pandemic (Caputo & Reichert 2020).

However, most of the current evidence points out to a decline in PA practice during this period, due to social distancing (Caputo & Reichert 2020). In Brazil, the early stages of social restriction were marked by a decline in PA practice and increase in screen time, ultra-processed food intake, cigarettes smoking, and alcohol consumption (Malta et al. 2020). This scenario affected the overall population, but the pattern was even more markedly observed among disadvantaged populations (Crochemore-Silva et al. 2020), since the political misconduct of the Brazilian government during the COVID-19 crisis led to the exacerbation of the already established social inequalities in the country (Ribeiro & Leist 2020; Hallal 2020).

Another group that could be heavily affected by the pandemic are university students and staff. To minimize the virus spread, Brazilian universities have been closed since March 2020 until mid 2022, which clearly impacted on the routine and behaviors of the academic community. Evidence from other countries suggests that COVID-19 might impact on sleep quality and mental health in this group (Kaparounaki et al. 2020; Wright et al. 2020; Marelli et al. 2021). However, there is still lack of evidence on the effects of the COVID-19 pandemic on the health-related behaviors of the Brazilian university community. Thus, the aim of the present study is to describe physical activity practice during the COVID-19 pandemic among students and staff of a southern Brazilian university, as well as its association with sociodemographic characteristics.

## Materials and Methods

This web-based cross-sectional study was carried out between July and August of 2020 in Pelotas, a city with approximately 340,000 inhabitants in Southern Brazil,(IBGE, 2020) and participants were from the Federal University of Pelotas community. We considered eligible all students (18,814 undergraduate and 3,781 graduate) and staff (1,369 academic staff and 1,256 administrative staff) of this public university. Each participant that accepted to participate, signed an informed consent term electronically before starting the survey. This study was approved by the Human Research Ethics Committee of the Federal University of Pelotas under protocol number 4.103.085.

Prior to data collection, the questionnaire was piloted in order to assess the understanding of the instruments used and its duration. All eligible participants received an email through the University system with information about the survey and the questionnaire link to take part on it. The questionnaire link was also made available on the survey official social media page on Instagram and on Facebook. The questionnaire was hosted online in the REDCap software (Harris et al. 2009). The first page of the questionnaire contained the informed consent form. To access the questionnaire, participants had to click ‘Yes’ after the question that asked whether they agreed to participate. If the participant agreed to participate, 65 mandatory close-ended items opened in the screen.

The main outcome for this study was leisure time physical activity (LTPA) during the COVID-19 pandemic measured by an adapted version of the leisure-time domain of IPAQ (International Physical Activity questionnaire), which measures frequency and intensity. We considered the individual as active if reporting ≥150 minutes of LTPA in the seven days prior to the survey. Two secondary outcomes were considered: first, if the individual practiced any physical activity (e.g.: gymnastics, sports, walking, running or other) before the pandemic and second, if the individual practiced any physical activity during the pandemic. In addition, for those individuals who performed any LTPA during the pandemic, we asked the participants about where they performed those physical activities (at home, indoor, outdoor, or other), if the participant received guidance by a trained professional (yes or no) – if yes, whether it was the same as before the COVID-19 pandemic, by new professional guidance, by videos or internet posts without professional bond, or other way. Lastly, we asked if the participants used to practice LTPA before social distancing started in Brazil.

The covariates used in this study were sex (male or female), age (<25; 25 – 34; ≥35 years for students and <35; 35 – 44; and ≥45 years for staff), family income in the last month (up to BRL 1,000; BRL 1,001 –2,500; BRL 2,501 – 5,000; BRL 5,001 – 10,000; more than BRL 10,001 for students and up to 5,000; BRL 5,001 – 10,000; more than BRL 10,001 for staff – note that US$1.00 was around BRL5.20 during data collection), self-reported skin color (white; black or brown; yellow or indigenous), household composition (live alone; live with family members; live with friends; live with a partner; other), maternal education (incomplete high school, complete high school, incomplete college, complete college, complete post-graduation), level of education (up to complete high school, incomplete college, complete college or complete post-graduation for students and incomplete college, complete college or complete post-graduation for staff), university position (undergraduate students, or graduate students/post-doc for students; administrative staff or academic staff for professional services staff), if the participant is currently employed (no; yes, up to 20h/week; yes, 20h-40h/week; or yes, more than 40h/week) – only for students –, and self-reported level of social distancing (low, some or quite hard/isolated).

The statistical analyses presented in this study were mainly descriptive, based on proportions and their respective 95% confidence intervals. Fisher exact test was used to evaluate the association between each of the covariates and the outcome. All analyses were performed in Stata version 15.1 (Stata Corporation, College Station, TX, USA), adopting a significance level of 5%.

## Results

The sample size comprised 2,853 individuals, of which 2,175 were students from the university and 678 were university staff, representing a response rate of 25.8% among students and 9.6% among university staff. Comparisons between the total university community and the studied sample according to gender, skin color and age is described in Supplementary Table I. According to Table I, most participants were female (71.0% for students and 60.6% for staff), white (78.3% for students and 92.9% for staff), lived with family members (59.8% for students and 45.8% for staff), and whose mothers did not complete high school (34.0% for students and 42.9% for staff). Considering only students, most of the sample was 25 years or younger (54.6%) and with a family income between BRL 1,001 – 2,500. Yet, considering only staff, we had a greater number of people with 45 years or older (47.3%), and earning more than BRL 10,000 (55.5%).

**Table I.**
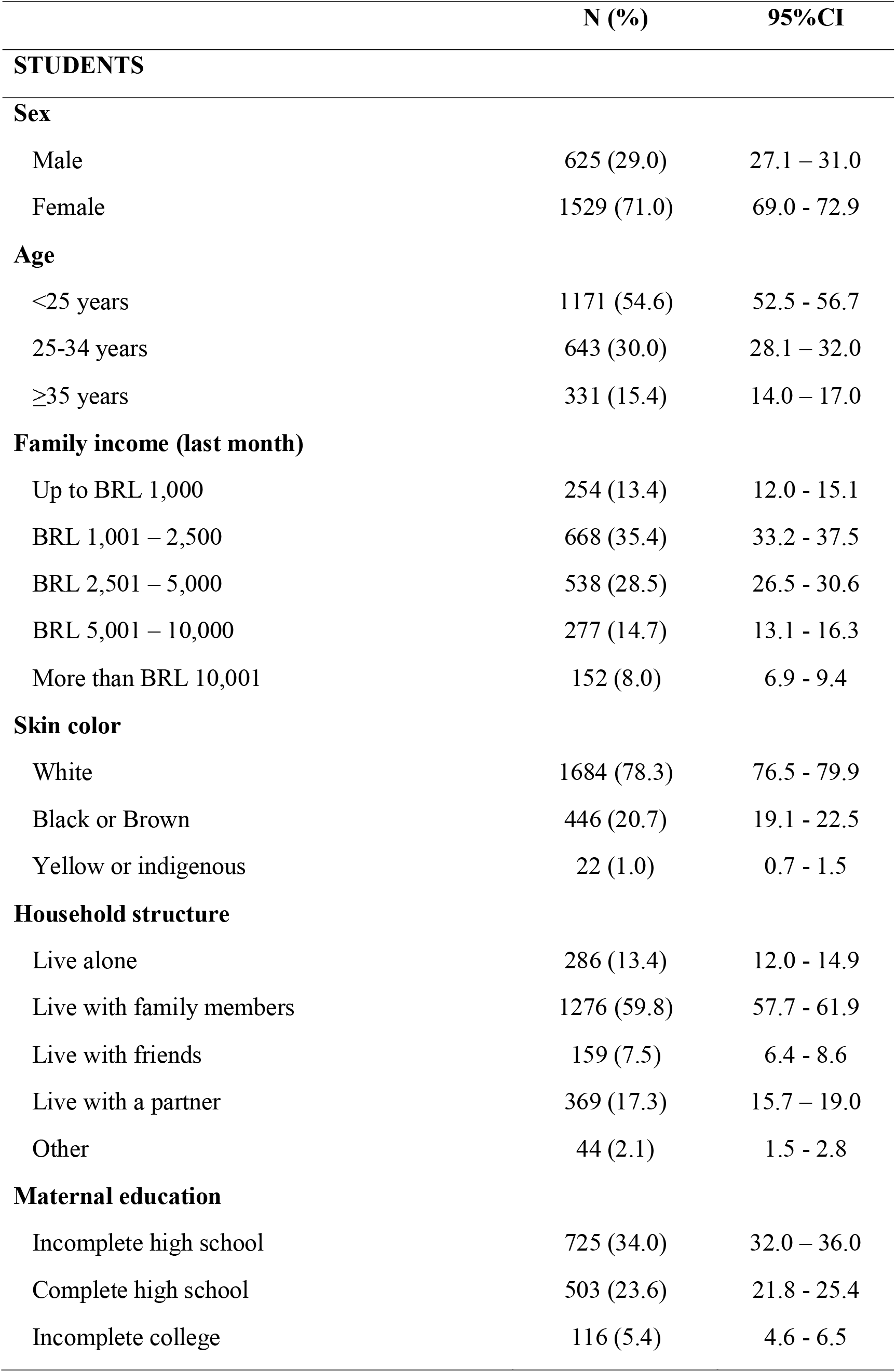
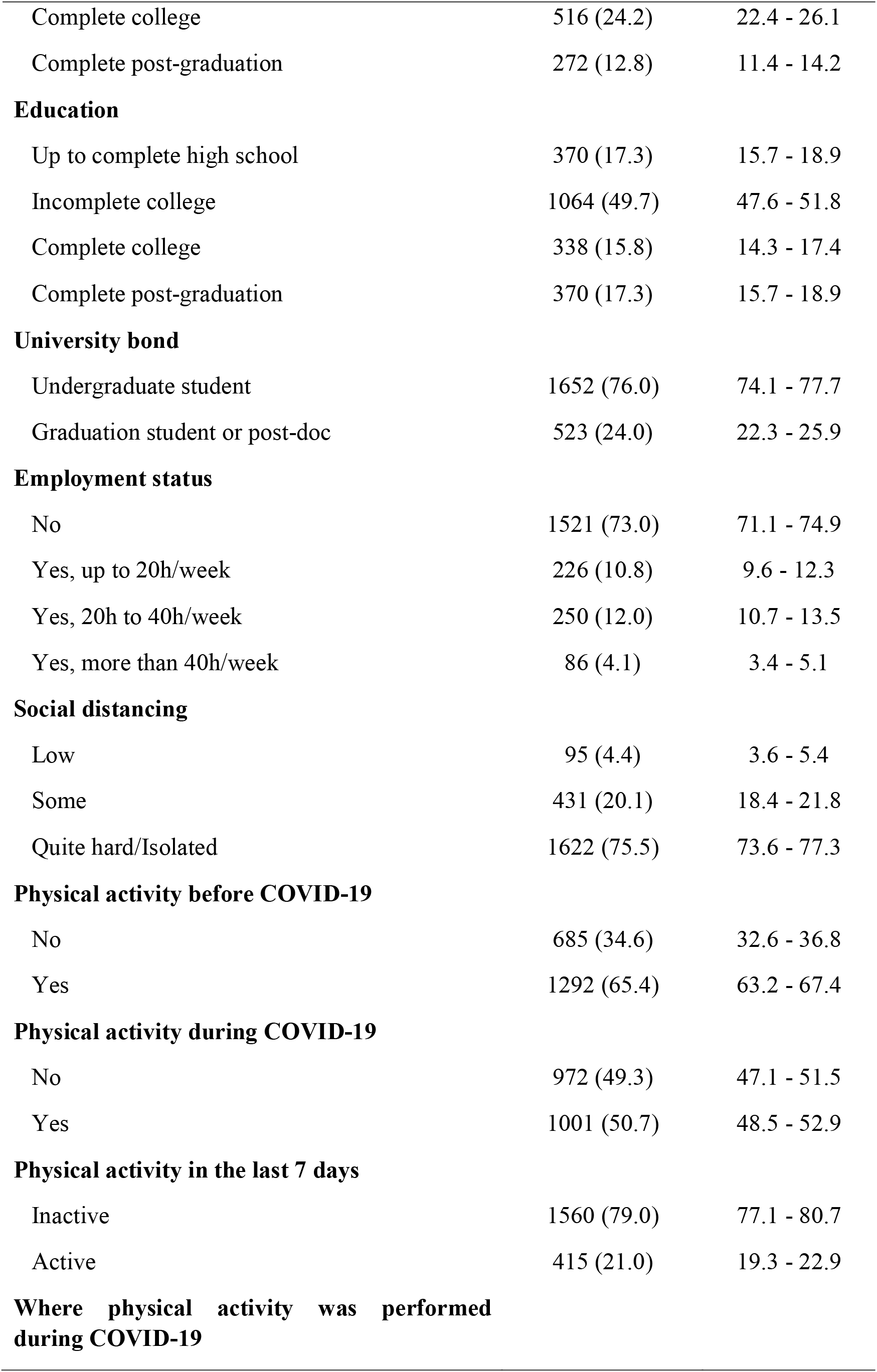
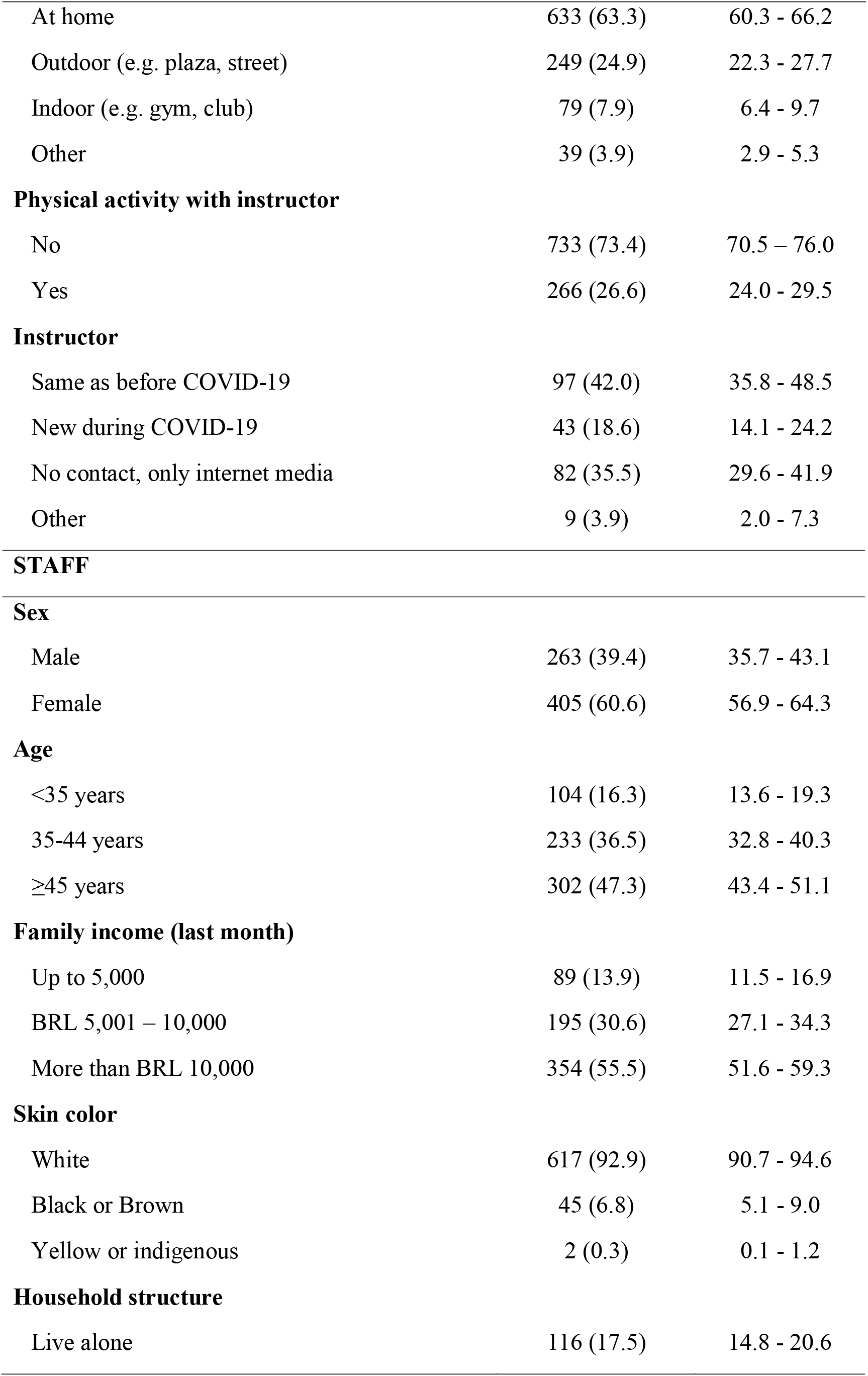
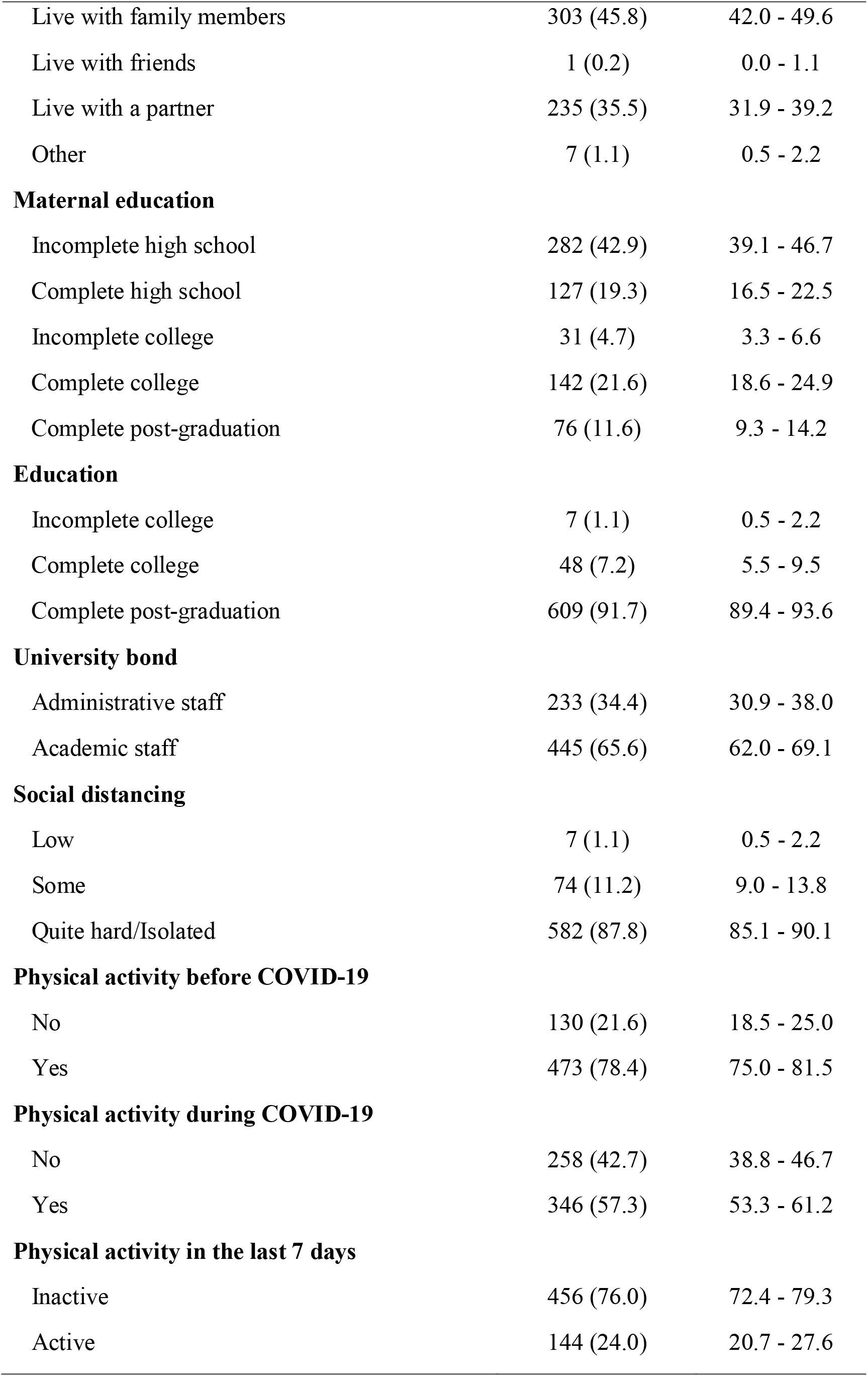
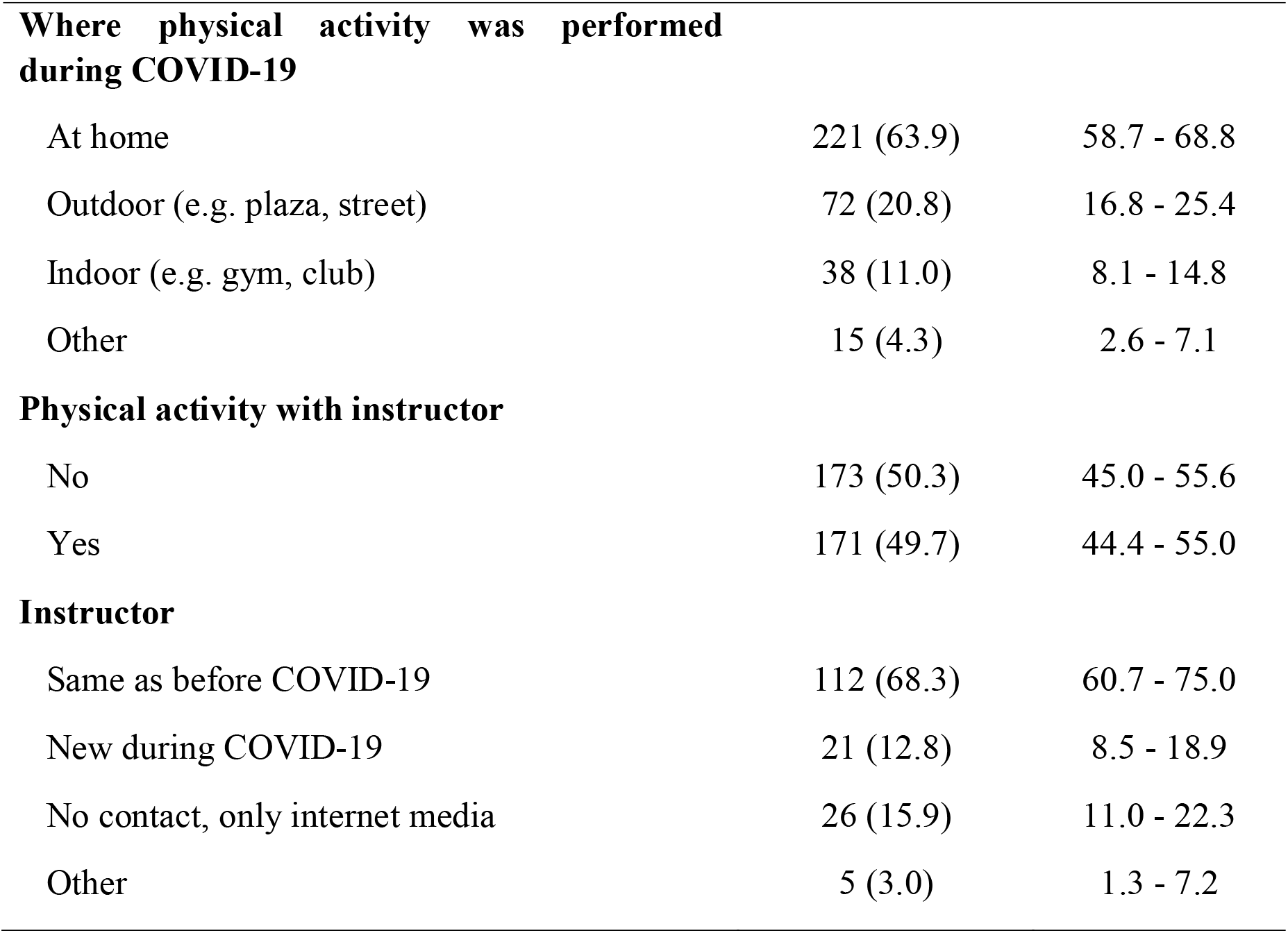
Descriptive characteristics of the analytic sample.

|  | **N (%)** | **95%CI** |
| --- | --- | --- |
| **STUDENTS** |  |  |
| **Sex** |  |  |
| Male | 625 (29.0) | 27.1 – 31.0 |
| Female | 1529 (71.0) | 69.0 - 72.9 |
| **Age** |  |  |
| <25 years | 1171 (54.6) | 52.5 - 56.7 |
| 25-34 years | 643 (30.0) | 28.1 – 32.0 |
| ≥35 years | 331 (15.4) | 14.0 – 17.0 |
| **Family income (last month)** |  |  |
| Up to BRL 1,000 | 254 (13.4) | 12.0 - 15.1 |
| BRL 1,001 – 2,500 | 668 (35.4) | 33.2 - 37.5 |
| BRL 2,501 – 5,000 | 538 (28.5) | 26.5 - 30.6 |
| BRL 5,001 – 10,000 | 277 (14.7) | 13.1 - 16.3 |
| More than BRL 10,001 | 152 (8.0) | 6.9 - 9.4 |
| **Skin color** |  |  |
| White | 1684 (78.3) | 76.5 - 79.9 |
| Black or Brown | 446 (20.7) | 19.1 - 22.5 |
| Yellow or indigenous | 22 (1.0) | 0.7 - 1.5 |
| **Household structure** |  |  |
| Live alone | 286 (13.4) | 12.0 - 14.9 |
| Live with family members | 1276 (59.8) | 57.7 - 61.9 |
| Live with friends | 159 (7.5) | 6.4 - 8.6 |
| Live with a partner | 369 (17.3) | 15.7 – 19.0 |
| Other | 44 (2.1) | 1.5 - 2.8 |
| **Maternal education** |  |  |
| Incomplete high school | 725 (34.0) | 32.0 – 36.0 |
| Complete high school | 503 (23.6) | 21.8 - 25.4 |
| Incomplete college | 116 (5.4) | 4.6 - 6.5 |
| Complete college | 516 (24.2) | 22.4 - 26.1 |
| Complete post-graduation | 272 (12.8) | 11.4 - 14.2 |
| **Education** |  |  |
| Up to complete high school | 370 (17.3) | 15.7 - 18.9 |
| Incomplete college | 1064 (49.7) | 47.6 - 51.8 |
| Complete college | 338 (15.8) | 14.3 - 17.4 |
| Complete post-graduation | 370 (17.3) | 15.7 - 18.9 |
| **University bond** |  |  |
| Undergraduate student | 1652 (76.0) | 74.1 - 77.7 |
| Graduation student or post-doc | 523 (24.0) | 22.3 - 25.9 |
| **Employment status** |  |  |
| No | 1521 (73.0) | 71.1 - 74.9 |
| Yes, up to 20h/week | 226 (10.8) | 9.6 - 12.3 |
| Yes, 20h to 40h/week | 250 (12.0) | 10.7 - 13.5 |
| Yes, more than 40h/week | 86 (4.1) | 3.4 - 5.1 |
| **Social distancing** |  |  |
| Low | 95 (4.4) | 3.6 - 5.4 |
| Some | 431 (20.1) | 18.4 - 21.8 |
| Quite hard/Isolated | 1622 (75.5) | 73.6 - 77.3 |
| **Physical activity before COVID-19** |  |  |
| No | 685 (34.6) | 32.6 - 36.8 |
| Yes | 1292 (65.4) | 63.2 - 67.4 |
| **Physical activity during COVID-19** |  |  |
| No | 972 (49.3) | 47.1 - 51.5 |
| Yes | 1001 (50.7) | 48.5 - 52.9 |
| **Physical activity in the last 7 days** |  |  |
| Inactive | 1560 (79.0) | 77.1 - 80.7 |
| Active | 415 (21.0) | 19.3 - 22.9 |
| **Where physical activity was performed during COVID-19** |  |  |
| At home | 633 (63.3) | 60.3 - 66.2 |
| Outdoor (e.g. plaza, street) | 249 (24.9) | 22.3 - 27.7 |
| Indoor (e.g. gym, club) | 79 (7.9) | 6.4 - 9.7 |
| Other | 39 (3.9) | 2.9 - 5.3 |
| **Physical activity with instructor** |  |  |
| No | 733 (73.4) | 70.5 – 76.0 |
| Yes | 266 (26.6) | 24.0 - 29.5 |
| **Instructor** |  |  |
| Same as before COVID-19 | 97 (42.0) | 35.8 - 48.5 |
| New during COVID-19 | 43 (18.6) | 14.1 - 24.2 |
| No contact, only internet media | 82 (35.5) | 29.6 - 41.9 |
| Other | 9 (3.9) | 2.0 - 7.3 |
| **STAFF** |  |  |
| **Sex** |  |  |
| Male | 263 (39.4) | 35.7 - 43.1 |
| Female | 405 (60.6) | 56.9 - 64.3 |
| **Age** |  |  |
| <35 years | 104 (16.3) | 13.6 - 19.3 |
| 35-44 years | 233 (36.5) | 32.8 - 40.3 |
| ≥45 years | 302 (47.3) | 43.4 - 51.1 |
| **Family income (last month)** |  |  |
| Up to 5,000 | 89 (13.9) | 11.5 - 16.9 |
| BRL 5,001 – 10,000 | 195 (30.6) | 27.1 - 34.3 |
| More than BRL 10,000 | 354 (55.5) | 51.6 - 59.3 |
| **Skin color** |  |  |
| White | 617 (92.9) | 90.7 - 94.6 |
| Black or Brown | 45 (6.8) | 5.1 - 9.0 |
| Yellow or indigenous | 2 (0.3) | 0.1 - 1.2 |
| **Household structure** |  |  |
| Live alone | 116 (17.5) | 14.8 - 20.6 |
| Live with family members | 303 (45.8) | 42.0 - 49.6 |
| Live with friends | 1 (0.2) | 0.0 - 1.1 |
| Live with a partner | 235 (35.5) | 31.9 - 39.2 |
| Other | 7 (1.1) | 0.5 - 2.2 |
| **Maternal education** |  |  |
| Incomplete high school | 282 (42.9) | 39.1 - 46.7 |
| Complete high school | 127 (19.3) | 16.5 - 22.5 |
| Incomplete college | 31 (4.7) | 3.3 - 6.6 |
| Complete college | 142 (21.6) | 18.6 - 24.9 |
| Complete post-graduation | 76 (11.6) | 9.3 - 14.2 |
| **Education** |  |  |
| Incomplete college | 7 (1.1) | 0.5 - 2.2 |
| Complete college | 48 (7.2) | 5.5 - 9.5 |
| Complete post-graduation | 609 (91.7) | 89.4 - 93.6 |
| **University bond** |  |  |
| Administrative staff | 233 (34.4) | 30.9 - 38.0 |
| Academic staff | 445 (65.6) | 62.0 - 69.1 |
| **Social distancing** |  |  |
| Low | 7 (1.1) | 0.5 - 2.2 |
| Some | 74 (11.2) | 9.0 - 13.8 |
| Quite hard/Isolated | 582 (87.8) | 85.1 - 90.1 |
| **Physical activity before COVID-19** |  |  |
| No | 130 (21.6) | 18.5 - 25.0 |
| Yes | 473 (78.4) | 75.0 - 81.5 |
| **Physical activity during COVID-19** |  |  |
| No | 258 (42.7) | 38.8 - 46.7 |
| Yes | 346 (57.3) | 53.3 - 61.2 |
| **Physical activity in the last 7 days** |  |  |
| Inactive | 456 (76.0) | 72.4 - 79.3 |
| Active | 144 (24.0) | 20.7 - 27.6 |
| **Where physical activity was performed during COVID-19** |  |  |
| At home | 221 (63.9) | 58.7 - 68.8 |
| Outdoor (e.g. plaza, street) | 72 (20.8) | 16.8 - 25.4 |
| Indoor (e.g. gym, club) | 38 (11.0) | 8.1 - 14.8 |
| Other | 15 (4.3) | 2.6 - 7.1 |
| **Physical activity with instructor** |  |  |
| No | 173 (50.3) | 45.0 - 55.6 |
| Yes | 171 (49.7) | 44.4 - 55.0 |
| **Instructor** |  |  |
| Same as before COVID-19 | 112 (68.3) | 60.7 - 75.0 |
| New during COVID-19 | 21 (12.8) | 8.5 - 18.9 |
| No contact, only internet media | 26 (15.9) | 11.0 - 22.3 |
| Other | 5 (3.0) | 1.3 - 7.2 |

According to Table II, for students, the prevalence of LTPA before and during the pandemic was 65.4% (70.0% for males and 63.5% for females) and 50.7% (52.2% for males and 50.1% for females), respectively. The prevalence of active individuals for the entire student sample was 21.0% (26.5% for males and 18.8% for females). For staff, the prevalence of LTPA before and during the pandemic was 78.4% (78.0% for males and 78.8% for females) and 57.3% (57.6% for males and 57.1% for females), respectively. The prevalence of active individuals for the entire staff sample was 24.0% (22.2% for males and 26.8% for females).

**Table II.** Prevalence of individuals practicing any LTPA before and during the pandemic, and prevalence of active individuals (≥150 minutes per week) considering only LTPA during the pandemic stratified by sex.

| **Students** |  |  |  |
| --- | --- | --- | --- |
|  | **Before COVID-19** | **During COVID-19** | **Active individuals during COVID-19** |
| Males | 70.0 (66.1 – 73.6) | 52.2 (48.1 – 56.3) | 26.5 (23.0 – 30.3) |
| Females | 63.5 (60.9 – 66.0) | 50.1 (47.5 – 52.8) | 18.8 (16.8 – 20.9) |
| Total | 65.4 (63.2 – 67.4) | 50.7 (48.5 – 52.9) | 21.0 (19.3 – 22.9) |
| **Staff** |  |  |  |
|  | **Before COVID-19** | **During COVID-19** | **Active individuals during COVID-19** |
| Males | 78.0 (72.2 – 82.8) | 57.6 (51.2 – 63.8) | 26.8 (21.5 – 32.9) |
| Females | 78.8 (74.3 – 82.6) | 57.1 (51.9 – 62.0) | 22.2 (18.2 – 26.8) |
| Total | 78.4 (75.0.- 81.6) | 57.3 (53.3 – 61.2) | 24.0 (20.7 – 27.6) |

According to Table III, the most active student males were those with brown or black skin color, those active before the pandemic and those practicing physical activity with an instructor. On the other hand, the less active student males were those with white skin color and those with low level of social distancing. Considering female students, the richest, those active before the pandemic and those practicing physical activity with an instructor were more active than their counterparts. In the contrary, the less active female students were those with complete high school and those with low level of social distancing. Among staff, both men and women were more active if they were already practicing physical activity before the pandemic. In addition, men with low level of social distancing and the poorest women were less active (Table III).

**Table III.**
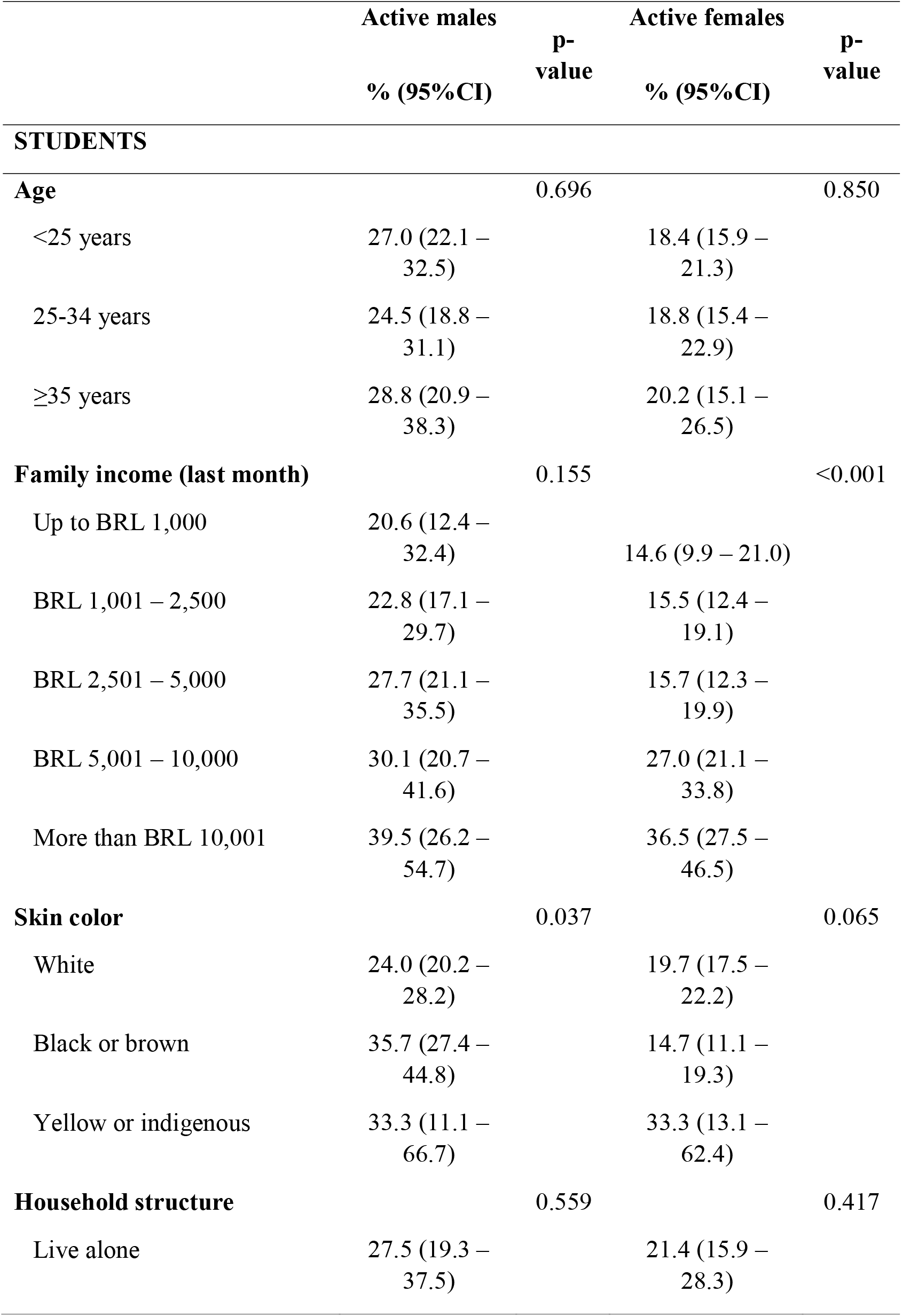
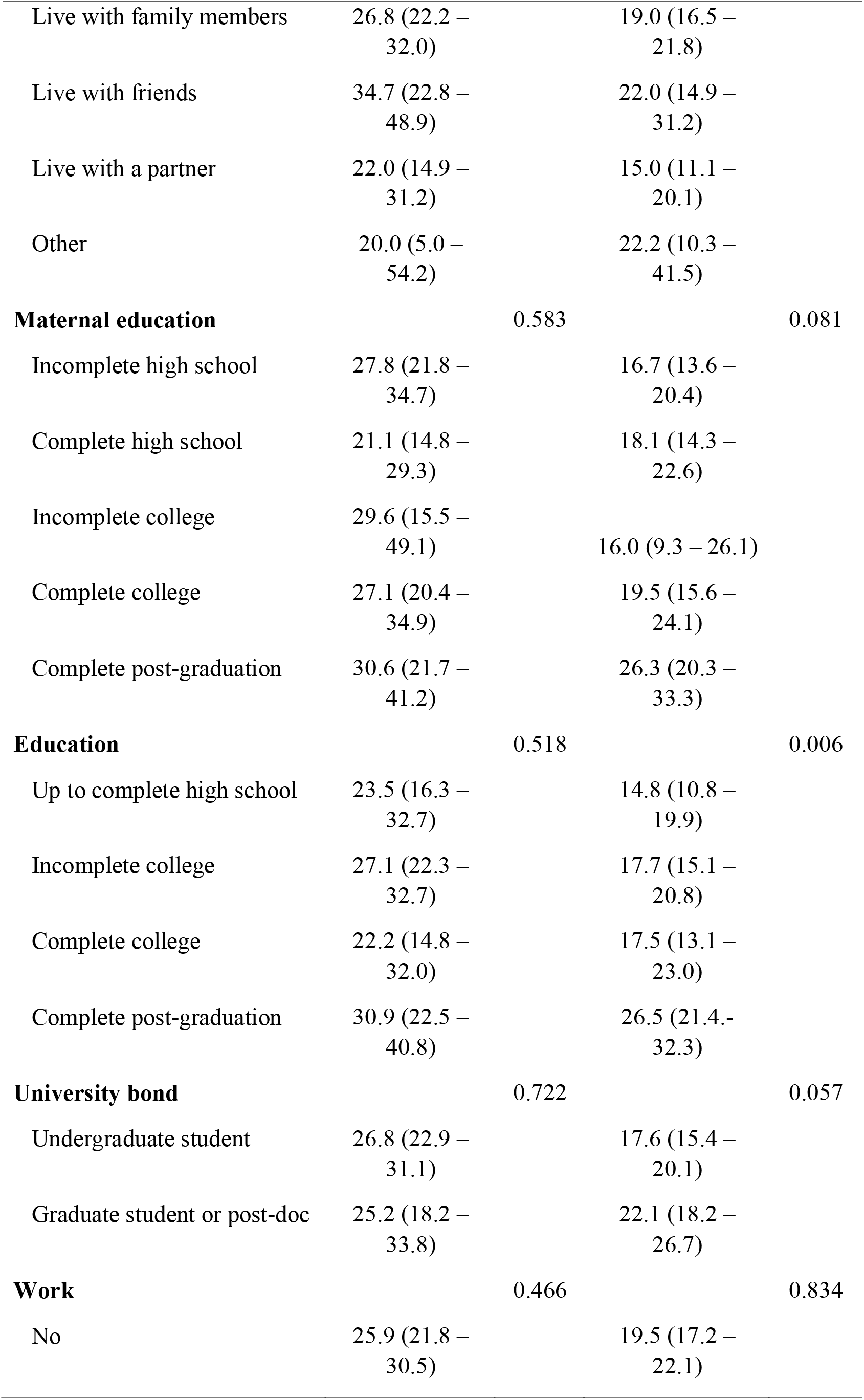
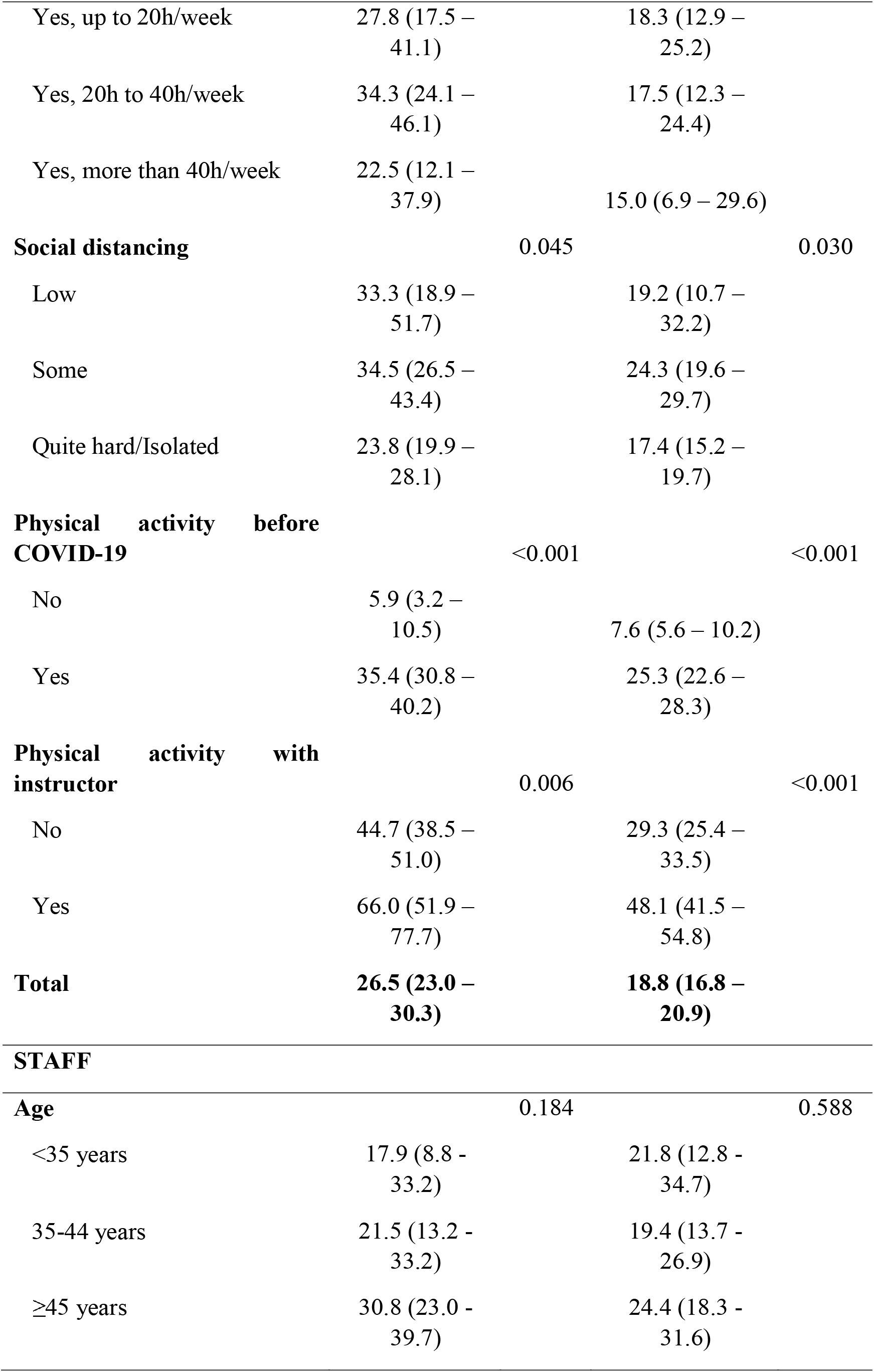
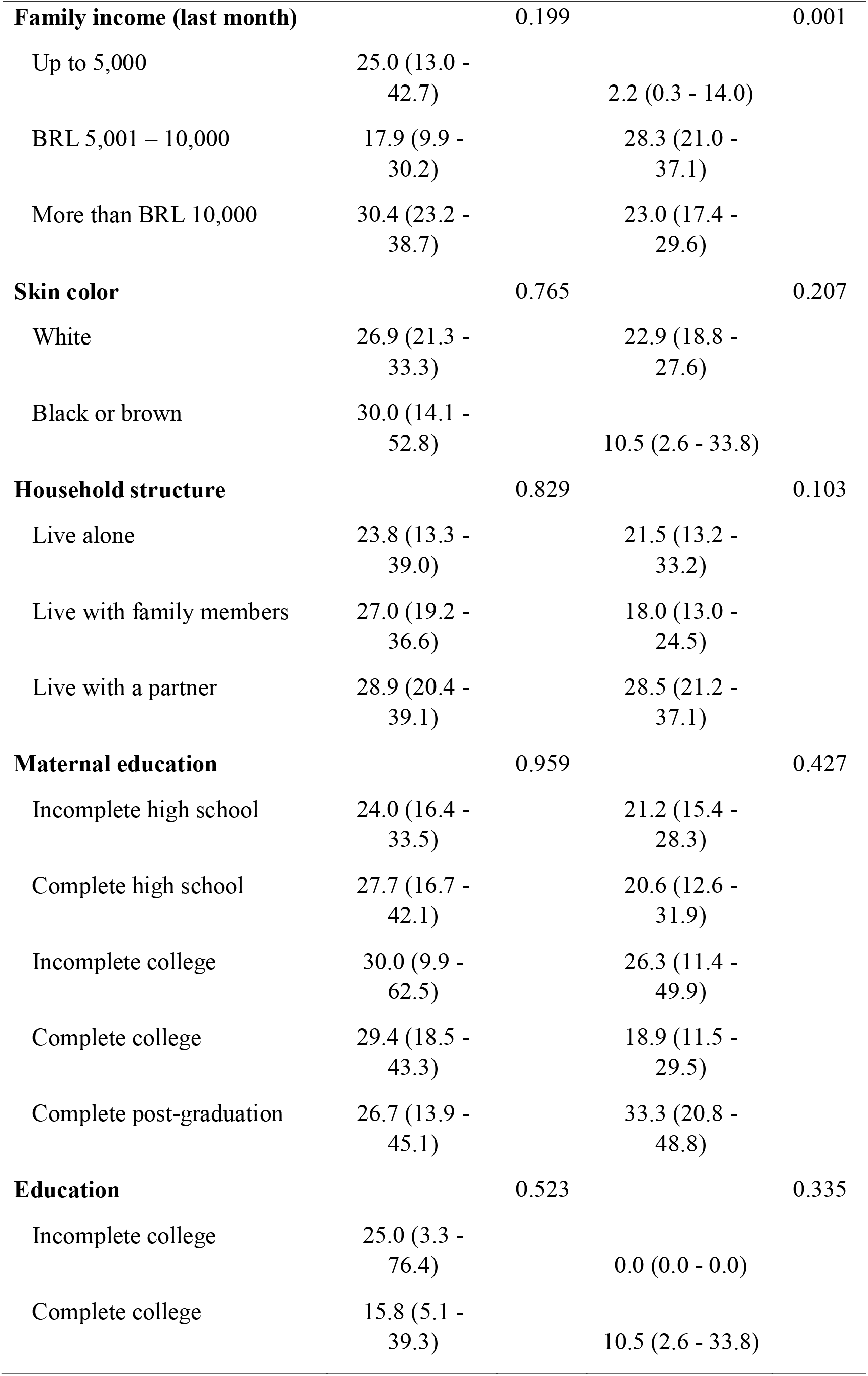
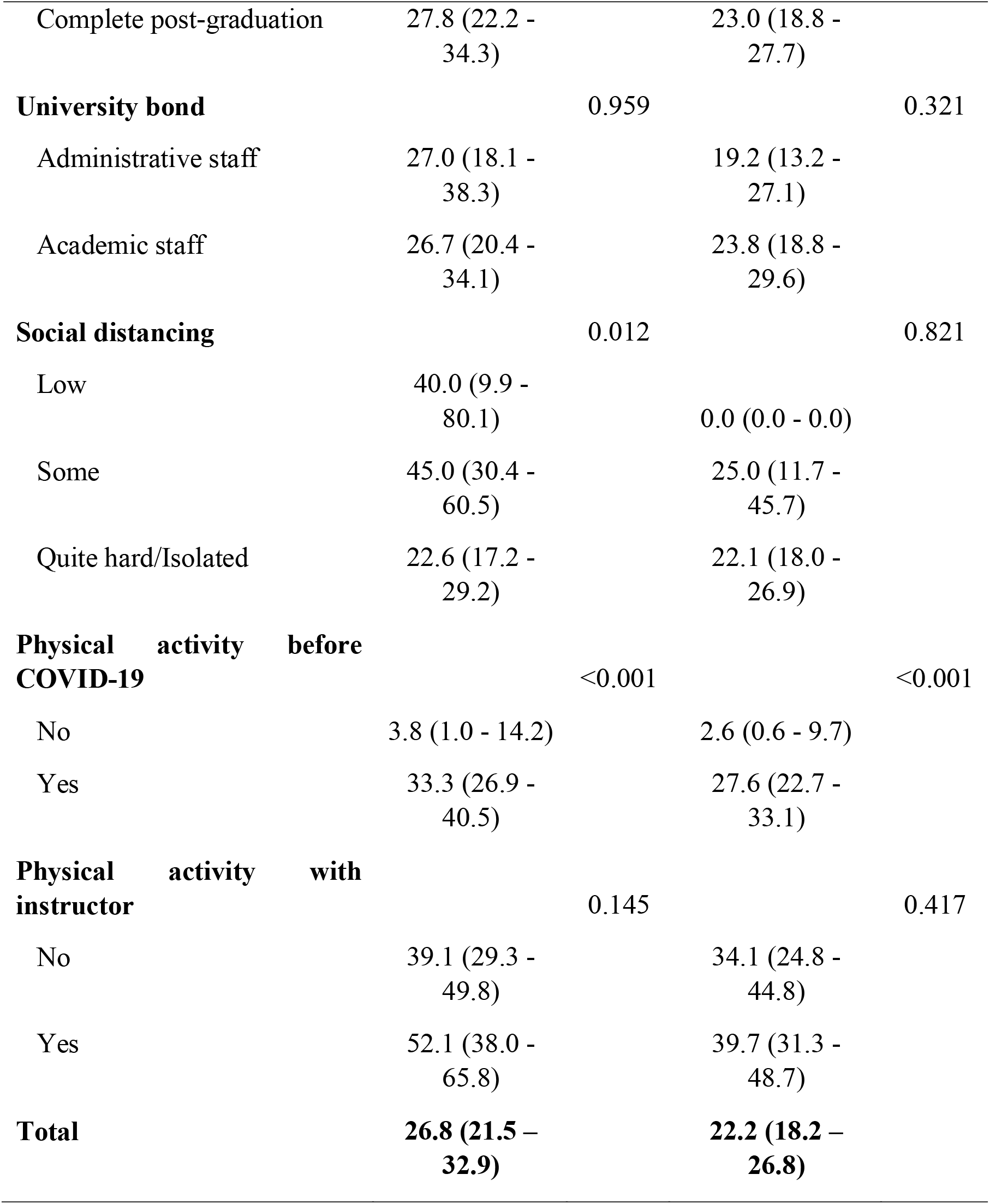
Physical activity (≥150 minutes in the last 7 days) during COVID-19 among students and sociodemographic variables.

Figure I shows the gender-specific prevalence of LTPA according to participant’s routine during COVID-19, stratified by students and staff. Participants that were following the protocols of staying at home were the ones with lower prevalence of LTPA. Among those participants reporting LTPA practice, most individuals practiced their physical activities at home, followed by outdoors (parks or plazas) for both males and females (Figure II). Lastly, Supplementary Figure I shows the prevalence of LTPA among active individuals according to professional aid. More than 50% of students and staff kept the same type of contact with trained professional as before the pandemic (either online or in person), and almost 20% of students and staff started online classes with physical education professionals.

**Figure I.**
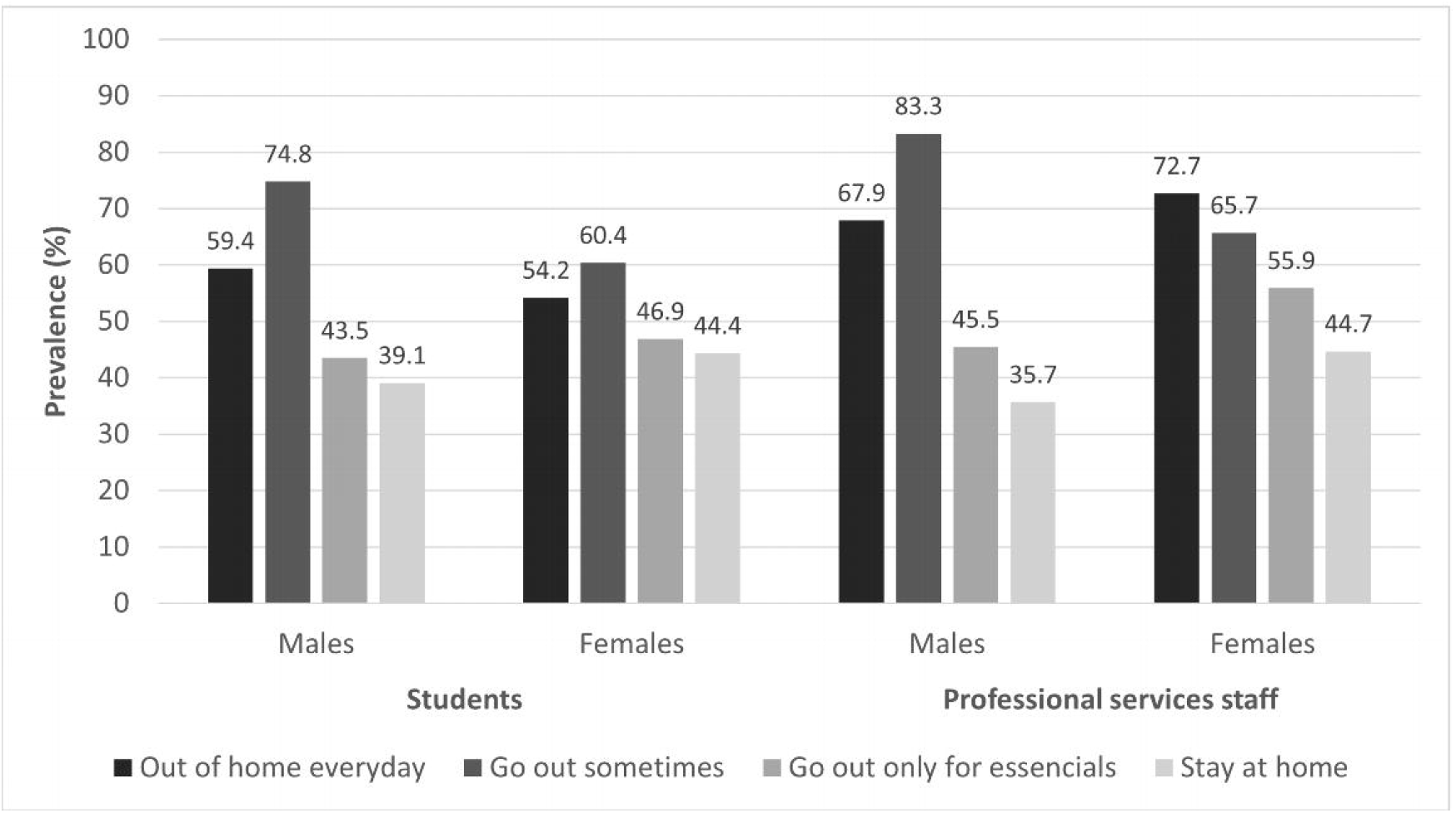
Gender-specific prevalence of LTPA according to participant’s routine during COVID-19

**Figure II.**
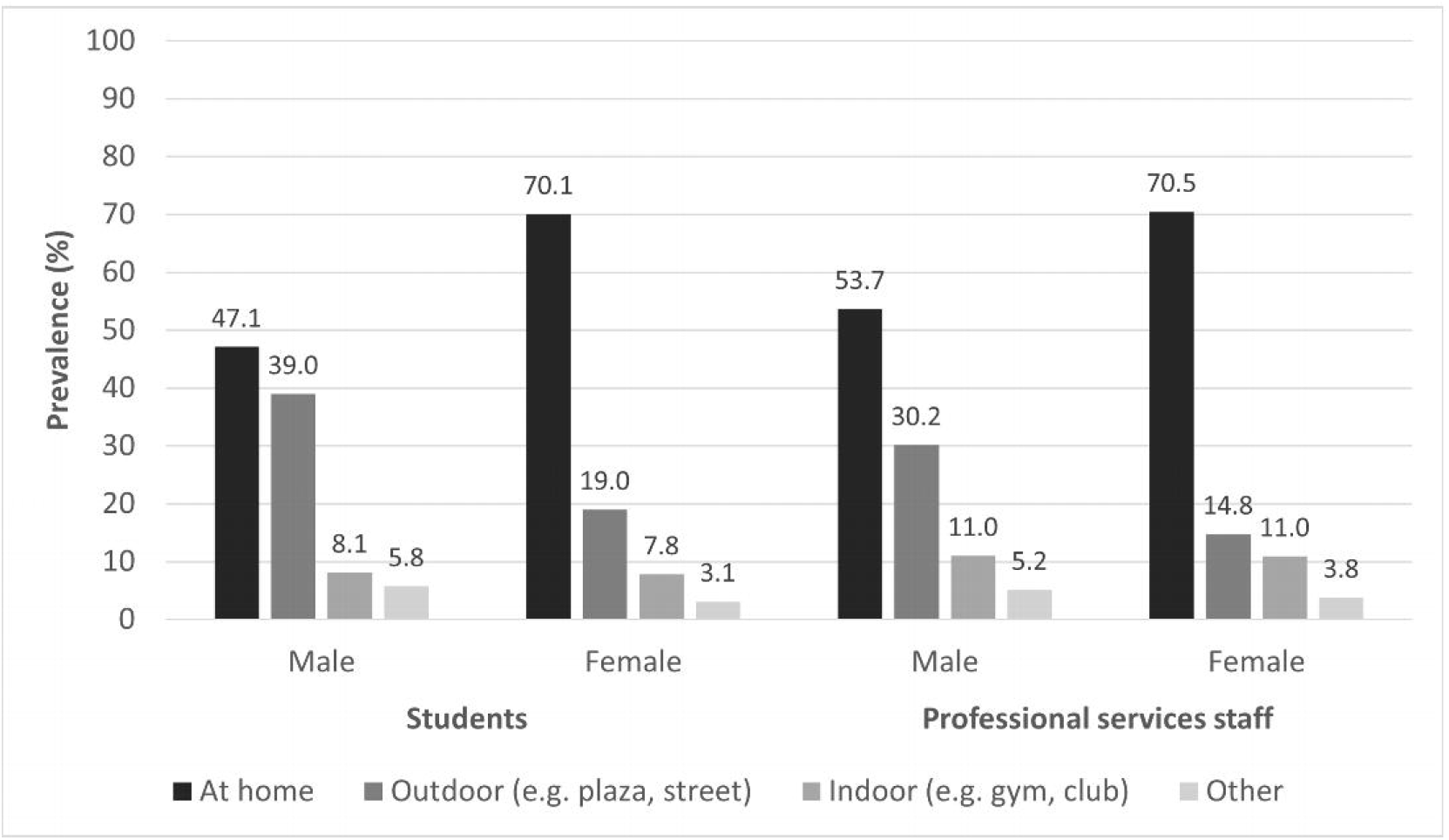
Gender-specific prevalence of LTPA according to participant’s place of practice

## Discussion

The present study described LTPA practices amid COVID-19 pandemic among students and staff of a southern Brazilian university, in addition to the association of the outcome with sociodemographic characteristics. Around one quarter of the participants reported achieving physical activity recommendations (≥150 minutes per week) only considering leisure-time. There was a decline of more than 15 percentage points in PA practice comparing pre- and during the pandemic, and those following the protocols of staying at home presented lower levels of LTPA. PA practice was mainly performed at home and without any professional help.

The pandemic has negatively impacted several outcomes of the world’s population, such as mental health (Loret de Mola et al. 2021; Sher 2020; Serafim et al. 2021; Schuch et al. 2021), food insecurity (Huizar et al. 2021), education (UNICEF, 2021), unemployment (Parolin 2020; Carvalho et al. 2021; Svabova et al. 2020), and sleep quality (Sher et al. 2020). In addition to that, the prevalence of physical activity has declined (Puccinelli et al. 2021). Considering university students’ samples, we found only one study that, surprisingly, had a positive effect of COVID-19 pandemic on physical activity levels (Romero-Blanco et al. 2020). All other studies identified declines in physical activity prevalence during the pandemic (Bertrand et al. 2021; Talapko et al. 2021; Gallo et al. 2020). A review of university students’ physical activity levels pre and during the pandemic also found a reduction on PA levels in 9 of the 10 studies included, in spite the overall low quality of evidence (López-Valenciano et al. 2021).

It is known that physical activity can improve the mental health burden of the world’s population (Chekroud et al. 2018). A study conducted in Japan analyzed the association of different domains of physical activity with risk of depressive symptoms among workers, and they found that while occupational and commuting physical activity did not change depressive symptoms, LTPA reduced the risk of depression (Kuwahara et al. 2015). At the same time, according to our data, just a few participants reported ≥150 minutes per week of leisure-time physical activity. Therefore, it is relevant to spread to the population the importance of maintaining a healthy routine (such as good diet and physical activity) for general wellbeing and immunity, as well as to help looking after a good mental health due to its impact during the pandemic.

Women’s family income impacted heavily on physical activity level both for students and staff, however this pattern was not observed for men. This could be due to the unequal gender labor distribution. It is still socially accepted that women, especially those with lower income, are the main caregiver of children and responsible for most of the household activities (Jolly et al. 2014). Summing this up to formal work, poorer women have been suffering from a double burden of activities for a long time. During the pandemic (with lockdowns and “stay at home” policies), that pattern increased gender inequalities, reducing women’s leisure time and, consequently, may decreased physical activity levels (Giordani et al. 2021; Wenham et al. 2020).

The reduction in PA practice is an expected development of the social distancing measures taken to reduce COVID-19 spread (Malta et al. 2020), since a favorable environment to practice PA at home is accessible to few. Our results showed that social distancing and a stay-at-home routine were related to lower prevalence of LTPA. Similarly, a systematic review study on PA and sedentary behavior from before to during the COVID-19 pandemic, found a decline in physical activity levels in 64 out of the 66 studies included, in which all of them reported a reduction higher than 50% (Stockwell et al. 2021).

Given the complex nature of PA, a wide set of contextual aspects can contribute to the choice or opportunity of practice. Online classes or personal trainers are usually expensive and require an adequate infrastructure (i.e., large room, silence, equipment) therefore, socioeconomically disadvantaged subgroups are less likely to maintain such activities during COVID-19 (Sallis & Pratt 2020). Also, it is possible that the level of priority of PA practice, or even other health-related aspects of life, was lowered, because of the economic challenges posed by the pandemic. The reduction in PA could be even more expressive among families with children or older people, in which the care for those individuals demands time and effort of the family, reducing the opportunity for LTPA.

On the other hand, our study demonstrates a positive impact of PA practice before the pandemic over the maintenance of this behavior during the COVID-19 social distancing period. We found no studies evaluating the effect of PA before the pandemic over the prevalence of PA during the pandemic. The literature is focused on PA changes during this period. Further research is needed to strengthen the evidence found in the present study.

Among participants reporting physical activity practice during pandemic, more than 50% among participants reporting LTPA practice during pandemic persisted doing classes or being supervised by physical education professionals as prior to the pandemic. Therefore, keeping contact with physical education professional might have impacted on the maintenance of physical activity behavior (Crochemore-Silva et al. 2020). Future studies may help to understand this relationship, that could also be influenced by the high socioeconomic profile, which both students and staff might have to be able to access private supervision for their physical activities, and to keep their routine stable even in a pandemic scenario.

Our study presents an important limitation regarding sample representativeness. Despite efforts for large participation of the university community, the representativeness of our sample is not guaranteed (Schuch et al. 2021). The analytical sample of our study is younger and presents a higher proportion of female individuals than the eligible university community, which could underestimate the physical activity prevalence found in the present study, since adult women usually present lower physical activity levels when compared to men (World Health Organization, 2015). On the other hand, the younger sample could have the opposite impact on our estimates (Sallis et al. 2016). Therefore, our results should be interpreted with caution. This limitation could be partially explained by the online nature of our research, which could negatively impact representativeness and response rate. In addition to sociodemographic characteristics, the students and staff who answered our questionnaire, compared to the non-participants, may have different relationship with the university, different levels of interest in the research topic might be affected in dissimilar proportions by the pandemic. In general, the university community is not representative of the entire population since those achieving university levels are already socioeconomically benefited individuals.

Nevertheless, the present study raises relevant descriptive insights on physical activity levels, their correlates and other characteristics of practice that can be important for institutions in the post-pandemic scenario. The LTPA prevalence during the pandemic was relatively low among students and staff and participants that followed staying at home protocols presented lower levels of physical activity.

## Data Availability

All data produced in the present study are available upon reasonable request to the authors

## Author contributions

**RCM:** conceptualization, methodology, formal analysis, writing original draft and final revision; **LICR:** writing original draft, formal analysis and final revision; **ICS:** conceptualization, writing original draft and final revision; **FFD:** investigation and final revision; **TNM:** investigation and final revision; **MLL:** investigation and final revision; **MGC:** investigation and final revision; **HSS:** investigation and final revision; **PCH:** supervision and final revision

## Figure legends

Supplementary Figure I. Prevalence of LTPA among active individuals according to professional aid

## Tables legends

Supplementary Table I. Comparison of analytic sample (n = 2,175 students and n = 678 staff) and eligible participants (n = 22,595 students and n = 2,625 staff)

