## Supplementary Table 1 for "How COVID-19 pandemic impacted the students and staff physical activity? A study in a Southern Brazilian University"

Supplementary Table 1. Comparison of analytic sample (n = 2,175 students and n = 678 staff) and eligible participants (n = 22,595 students and n = 2,625 staff)

|  | **Students** | | | | **Staff** | | | |
| --- | --- | --- | --- | --- | --- | --- | --- | --- |
|  | **UFPel** | | **Analytic sample** | | **UFPel** | | **Analytic sample** | |
|  | **N** | **%** | **N** | **%** | **N** | **%** | **N** | **%** |
| Sex | p<0.001 | | | | p=0.001 | | | |
| Male | 10,144 | 44.9 | 625 | 29.0 | 1,221 | 46.5 | 263 | 39.4 |
| Female | 12,451 | 55.1 | 1,529 | 71.0 | 1,404 | 53.5 | 405 | 60.6 |
| **Total** | **22,595** | **100.0** | **2,154** | **100.0** | **2,625** | **100.0** | **668** | **100.0** |
| Skin color | p=0.195 | | | | p=0.333 | | | |
| White | 14,266 | 76.9 | 1684 | 78.3 | 2,349 | 91.1 | 617 | 92.9 |
| Black or Brown | 4,124 | 22.2 | 446 | 20.7 | 219 | 8.5 | 45 | 6.8 |
| Yellow or Indigenous | 156 | 0.9 | 22 | 1.0 | 10 | 0.4 | 2 | 0.3 |
| **Total** | **18,546** | **100.0** | **2,150** | **100.0** | **2,578** | **100.0** | **664** | **100.0** |
| Age (in years) | p<0.001 | | | | p=0.009 | | | |
| <25 | 10,671 | 47.3 | 1,171 | 54.6 | 3 | 0.1 | 4 | 0.6 |
| 25-30 | 5,937 | 26.4 | 503 | 23.5 | 81 | 3.1 | 31 | 4.7 |
| ≥31 | 5,934 | 26.3 | 471 | 22 | 2,541 | 96.8 | 629 | 94.7 |
| **Total** | **22,542** | **100.0** | **2,145** | **100.0** | **2,625** | **100.0** | **664** | **100.0** |
