## Supplementary figures and images for "How COVID-19 pandemic impacted the students and staff physical activity? A study in a Southern Brazilian University"

### Supplementary Figure 1

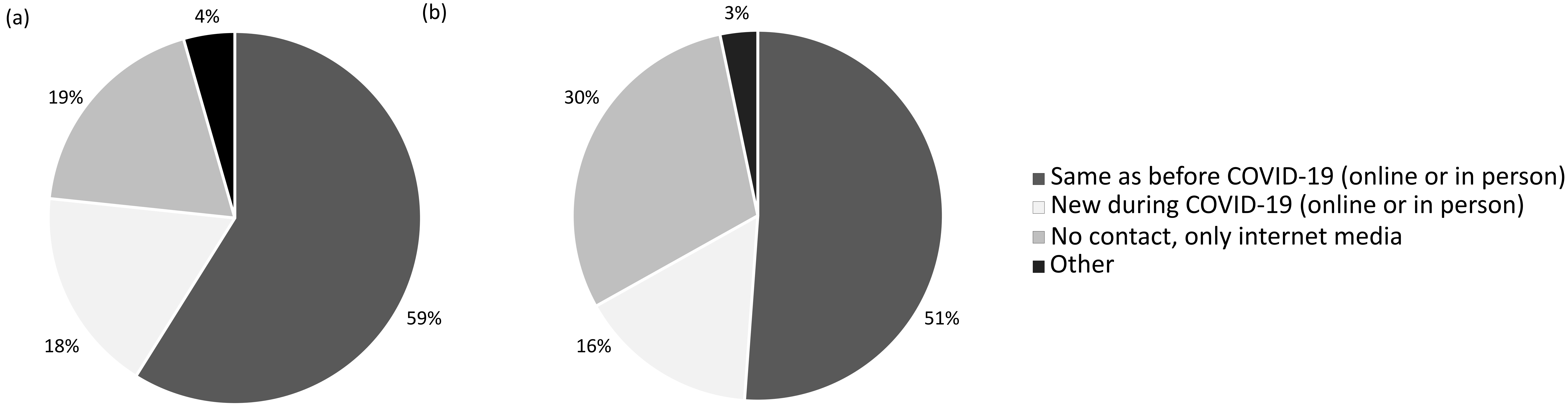
